# Finding the Clinical Traces of Cognitive Impairment in Patients with Heart Failure: A Natural Language Processing Study of Clinical Letters from Routine Care

**DOI:** 10.1101/2025.02.12.25322151

**Authors:** L. Malin Overmars, Bram van Es, Sander C. Tan, Jet M. J. Vonk, Majon Muller, Mark C. H. De Groot, Geert Jan Biessels, M.G. Van der Meer, Wouter W. van Solinge, Saskia Haitjema, Michiel L. Bots, Lieza G. Exalto

## Abstract

Cognitive impairment is common in patients with heart failure, but to which extent cognitive complaints are evaluated and listed in clinical practice is unknown. Therefore, this study aims to identify whether cognitive complaints are listed in clinical notes of patients with heart failure, consistent with listed complaints in clinical notes of patients attending memory clinics, by using natural language processing (NLP) techniques. Patients with heart failure and patients attending a memory outpatient clinic were identified by using echocardiography reports and presence of memory outpatient clinic codes stored in the Utrecht Individual-Oriented Database (UPOD) from 2011 to 2023. Named Entity Detection and Linking (NER+L) strategies MedCAT and MedCATTrainer were used to extract listed complaints in clinical notes by patient group, and it was assessed whether cognitive complaints were listed in clinical notes of patients with heart failure. Among 5803 patients with heart failure, dyspnea (57.1%), chest pain (48.4%), and oedema (43.6%) were the most listed complaints. In 967 patients attending memory clinics, memory problem (80.9%), getting lost (24.1%), and being morose (22.5%) were the most listed complaints. Notably, in patients with heart failure, the reporting of memory problems was low at 2.6%. This study shows a low reporting frequency of cognitive complaints in clinical notes of patients with heart failure, even though both conditions often co-occur according to cross-sectional studies. This points towards a potential underrecognition of cognitive complaints in patients with heart failure during clinical practice.

## Introduction

Cognitive impairment is common in patients with heart failure^1–4^. Both conditions are on the rise, particularly among the aging population in high-income countries^2,5^. Recognizing the coexistence cognitive impairment and of heart failure is crucial due to its significant clinical implications, as cognitive impairment can hinder medication adherence, self-care behaviours, and overall quality of life, while increasing the risk of adverse events in patients with heart failure^2,3,5–8^. However, the extent to which cognitive complaints are evaluated and listed in clinical notes of patients with heart failure in electronic health records (EHRs) is unknown.

The application of Natural Language Processing (NLP) to textual data stored in electronic health records (EHRs) has enabled the analysis of listed cognitive symptoms in patients with heart failure. While EHRs contain substantial structured data, such as laboratory results, blood pressure readings, and medication prescriptions, a considerable amount of clinical information is embedded in unstructured free text fields that have historically posed challenges in terms of analysis^9,10^. Recent advancements in NLP have demonstrated considerable potential in extracting pertinent data from such texts to address healthcare inquiries^9–13^. However, it should be noted that most NLP techniques have been developed for English-language medical texts, thereby constraining their applicability to other languages. The development of tools such as the Medical Concept Annotation Tool (MedCAT) and MedCATTrainer has rendered it feasible to automatically identify and link medical concepts, including patient complaints, to established Dutch biomedical ontologies^14–16^,. These advancements allow analysis of unstructured EHR data in Dutch and pave the way for research on symptom documentation.

This study aims to showcase the application of NER+L on Dutch clinical notes, by gaining real-world insight into the type and frequency of listed cognitive complaints in patients with heart failure, consistent with listed complaints in clinical notes of patients attending memory clinics, during routine clinical practice.

## Materials and Methods

### Data extraction and patient selection

Clinical notes were extracted from the Utrecht Patient-Oriented Database (UPOD) for visits at the University Medical Centre Utrecht (UMC Utrecht) between November 2011 and August 2023. We focused on two patient populations: those with heart failure and those attending memory clinics. Patients with heart failure were identified based on diagnostic codes recorded in UPOD, while memory clinic patients were selected using specific appointment codes (NGEH, NGEH⁺, NGEHP, NGEHV, and NVCI). Only correspondence documents labelled as either ‘*Poliklinische Brief’* or ‘*Klinische Brief’* and containing the keyword “Anamnese” were retained. Importantly, the analysis was conducted at the level of individual visits (unique letters) rather than unique patients.

### Text extraction and preprocessing

To isolate the relevant text, we first extracted the anamnesis sections from the full clinical reports using tailored regular expressions. These regex patterns were designed to accommodate variations in paragraph delimiters—including single or multiple newlines with or without intervening spaces—and common section headers.

\# Split the text into paragraphs using double newline as the delimiter

paragraphs <-unlist(strsplit(data$text, "\\r\\n\\r\\n"))

\# Keep only the paragraph that starts with "Anamnese"

anamnese_paragraph <-paragraphs[grep("^Anamnese", paragraphs)]

We then applied lemmatization using the NLTK library to convert words into their base forms (e.g., “running” became “run”), thereby standardizing the vocabulary for subsequent analysis.

### Model training and concept enrichment

To accurately extract and interpret complaints from the cleaned clinical notes, we utilized the MedCATTrainer and MedCAT libraries^14–16^,. A subset of 200 clinical notes was manually annotated. These manual annotations provided input for refining the MedCAT model and were used to enrich the underlying Dutch Medical Concept Database (CDB). The CDB initially comprised medical concepts from the 2022AB release of the Unified Medical Language Systems (UMLS; 254,835 concepts) and was subsequently expanded with additional terms and variants specific to our dataset. This enrichment was essential for improving the model’s accuracy, particularly for nuances in cognitive complaints.

### Named Entity Recognition and Entity Linking

MedCAT was then employed to perform Named Entity Recognition (NER) on the preprocessed text. The model leveraged 300-dimensional Word2Vec embeddings—trained on the MIMIC-III dataset—to capture contextual nuances, allowing for the accurate detection of complaint mentions even when synonyms, variations, or typographical errors were present. Following NER, MedCAT performed entity linking by mapping each identified complaint to a corresponding concept in the enriched CDB through a context-aware disambiguation process. For instance, when encountering an ambiguous term such as “confusion” (which could denote either a transient state or a sign of cognitive impairment), the system generated a set of candidate concepts. It then extracted the surrounding context and converted it into a 300-dimensional vector. By comparing this context vector to pre-stored context representations for each candidate using cosine similarity, MedCAT determined which concept best matched the context. If the surrounding text included indicators like “memory loss,” “disorientation,” or “cognitive decline,” the candidate representing cognitive impairment received a higher similarity score and was selected— provided the confidence score exceeded a predefined threshold.

### Filtering of relevant semantic types

After entity linking, recognized complaints were manually filtered to retain only those corresponding to semantic types relevant to our study. The selected semantic types included T184 (Sign or Symptom), T067 (Phenomenon or Process), T033 (Finding), T047 (Disease or Syndrome), T055 (Individual Behaviour), T041 (Mental Process), T054 (Social Behaviour), T039 (Physiologic Function), and T048 (Mental or Behavioural Dysfunction). This broader selection was necessary to capture conditions such as cognitive impairments that might not be adequately categorized under T184 alone. Additionally, we computed the length of each extracted anamnesis (in number of characters) and compared these lengths between memory clinic patients and patients with heart failure using appropriate statistical tests (with a significance threshold of p < 0.05).

### Association Rule Mining

To explore the relationships between complaints—particularly the co-occurrence of cognitive complaints with common heart failure–related complaints—we applied Association Rule Mining using the Apriori algorithm. This method generated if-then rules of the form {X ⇒ Y}, where {X} represents heart failure-related concepts (e.g., dyspnea, edema, or chest pain) and {Y} represents cognitive complaints (e.g., memory problems). The strength of these associations was quantified using a confidence measure, defined as the number of clinical notes containing both concept sets divided by the number of clinical notes containing the heart failure-related concepts.

### Software

The NLP-procedures were performed using Python version 3.10.7^17^ and R version 4.4.2^18^.

## Results

### Patient characteristics

Of patients seen at UMC Utrecht between November 2011 and August 2023, we analyzed 5803 clinical notes from patients with heart failure and 961 notes of patients attending memory clinics. The mean age of patients with heart failure was 61.3 years (SD = 14.8), with 1,887 (32.5%) being women, whereas memory clinic patients had a mean age of 67.0 years (SD = 12.7) and 454 (46.9%) were women. Notably, 47 patients (0.7%) were identified as having both heart failure and memory clinic attendance (Supplementary Figure S1). Additionally, clinical notes in letters for memory clinic patients were substantially longer, with a mean of 1,105.6 characters, compared to a mean of 518.9 characters for patients with heart failure (p < 0.001; Supplementary Figure S2).

### Commonly listed complaints in patients attending memory clinics

A total of 343 unique complaints were identified in the clinical notes of memory clinic patients. The top ten complaints for these patients included memory problem (n = 782, 80.9%), gets lost (n = 233, 24.1%), morose (n = 218, 22.5%), word finding disorder (n = 171, 17.7%), headache (n = 143, 14.7%), hallucination (n = 137, 14.2%), gets angry easily (n = 99, 10.2%), impaired concentration (n = 99, 10.2%), concentration normal (n = 95, 9.8%), and lack of energy (n = 80, 8.3%) (Figure 3).

### Commonly listed complaints in patients with heart failure

In patients with heart failure, 534 unique complaints were identified. The top ten complaints reported in this group were dyspnea (n = 3,312, 57.1%), chest pain (n = 2,808, 48.4%), oedema (n = 2,652, 45.7%), palpitations (n = 2,257, 38.9%), orthopnoea (n = 1,797, 30.1%), dizziness (n = 1,663, 28.7%), syncope (n = 1,194, 20.6%), pounding heart (n = 1,056, 18.3%), chest pressure (n = 824, 14.2%), and cough (n = 809, 13.9%) (Figure 2).

### Reporting of cognitive complaints in patients with heart failure

The top 25 complaints identified in patients attending memory clinics were compared with their reporting in patients with heart failure, stratified by sex (Table 1). In patients with heart failure, memory problems were reported in only 2.5% of cases overall (2.4% in women and 2.6% in men). Additional cognition-related complaints such as gets lost, morose, word finding disorder, hallucination, impaired concentration, gets angry easily, slow, mental orientation, and loss of initiative were each reported in less than 1.0% of patients with heart failure, irrespective of sex. In contrast, non-cognitive complaints such as dizziness (reported in 28.7% overall, 28.2% in women, and 28.8% in men), nausea (12.9% overall; 15.6% in women, 11.6% in men), and lack of energy (12.4% overall; 13.7% in women, 11.8% in men) were relatively common among patients with heart failure. These findings indicate that, except for dizziness, nausea, and lack of energy, the complaints frequently listed in memory clinic patients are rarely reported in patients with heart failure.

**Table 1.** Top-25 complaints listed in patients attending memory clinics and their reporting in patients with heart failure

| Listed complaint in patients attending memory clinics (Top-25) | Patients with heart failure (N =5803) | Women with heart failure (N = 1887) | Men with heart failure (N = 3916) |
| --- | --- | --- | --- |
| Memory problem (N, %) | 146 (2.6) | 45 (2.4) | 101 (2.6) |
| Gets lost (N, %) | 10 (< 1.0) | 2 (< 1.0) | 8 (< 1.0) |
| Morose (N, %) | 20 (< 1.0) | 5 (< 1.0) | 15 (< 1.0) |
| Word Finding Disorder (N, %) | 15 (< 1.0) | 1 (< 1.0) | 14 (< 1.0) |
| Headache (N, %) | 191 (3.3) | 65 (3.4) | 126 (< 1.0) |
| Hallucination (N, %) | 9 (<1.0) | 6 (< 1.0) | 3 (< 1.0) |
| Impaired concentration (N, %) | 15 (<1.0) | 4 (< 1.0) | 11 (< 1.0) |
| Gets angry easily (N, %) | 20 (<1.0) | 6 (< 1.0) | 14 (< 1.0) |
| Concentration normal (N, %) | 23 (<1.0) | 8 (< 1.0) | 15 (< 1.0) |
| Lack of energy (N, %) | 721 (12.4) | 259 (13.7) | 462 (11.8) |
| Slow (N, %) | 57 (<1.0) | 15 (< 1.0) | 42 (1.1) |
| Dizziness (N, %) | 1663 (28.7) | 532 (28.2) | 1131 (28.8) |
| Mental orientation (N, %) | 1 (<1.0) | 0 (0) | 1 (< 1.0) |
| Nausea (N, %) | 750 (12.9) | 295 (15.6) | 455 (11.6) |
| Loss of initiative (N, %) | 45 (<1.0) | 11 (< 1.0) | 34 (< 1.0) |
| Eerie feeling (N, %) | 98 (1.7) | 37 (1.9) | 61 (1.6) |
| Fear (N, %) | 88 (1.5) | 32 (1.7) | 56 (1.4) |
| Tired (N, %) | 528 (9.1) | 170 (9.0) | 358 (9.1) |
| Delusions (N, %) | 5 (<1.0) | 2 (< 1.0) | 3 (< 1.0) |
| Decreasing weight (N, %) | 468 (8.1) | 157 (8.3) | 311 (7.9) |
| Tension (N, %) | 90 (1.6) | 27 (2.2) | 63 (2.3) |
| Endogenous depressions (N, %) | 13 (< 1.0) | 3 (0.2) | 10 (0.3) |
| Dyspnea (N, %) | 3312 (57.1) | 1052 (55.7) | 2260 (57.7) |
| Stress (N, %) | 232 (4.0) | 96 (5.1) | 136 (3.5) |
| Difficulty sleeping (N, %) | 132 (2.3) | 48 (2.5) | 84 (2.1) |

### Association Rule Mining

We further examined the co-occurrence of cognitive complaints with common heart failure–related complaints using the Apriori algorithm for Association Rule Mining. For example, the rule {dyspnea} ⇒ {memory problem} suggests that when dyspnea is present in an anamnesis, the probability of also reporting a memory problem is 0.030; this co-occurrence was observed in 98 out of 5,803 patients with heart failure. Similarly, the rules {oedema} ⇒ {memory problem} and {chest pain} ⇒ {memory problem} had confidence values of 0.031 (observed in 82 patients) and 0.028 (observed in 78 patients), respectively. Additional association rules are detailed in Table 2 and Supplementary Table S2. Overall, these results demonstrate that the complaints commonly observed in memory clinic patients rarely co-occur with the typical complaints reported by patients with heart failure.

**Table 2.** Results of Apriori algorithm to assess co-occurrence of commonly listed complaints in patients attending at memory clinics in clinical notes of patients with heart failure.

| LHS |  | RHS | Confidence | Count (# clinical notes) |
| --- | --- | --- | --- | --- |
| {Dyspnea} | => | {Memory Problem} | 0.03 | 98 |
| {Hydrops} | => | {Memory Problem} | 0.03 | 82 |
| {Chest Pain} | => | {Memory Problem} | 0.03 | 78 |
| {Palpitations} | => | {Memory Problem} | 0.03 | 65 |
| {Dyspnea, Hydrops} | => | {Memory Problem} | 0.03 | 64 |
| {Chest Pain, Dyspnea} | => | {Memory Problem} | 0.03 | 63 |
| {Dizziness} | => | {Memory Problem} | 0.04 | 62 |

LHS = Left-Hand Side, RHS = Right-Hand Side, Confidence = The probability that a complaint Y (RHS) is listed given that complaint X (LHS) is listed (*for example, given that Dyspnea is listed, the probability that Memory Problem is listed is 0.03*).

## Discussion

This study demonstrates that complaints commonly reported in patients attending memory clinics—such as memory problems and word-finding difficulties—are infrequently documented in the clinical letters of patients with heart failure. Except for non-specific symptoms like dizziness, nausea, and lack of energy, cognitive complaints appear markedly underreported. In our dataset, only 2.5% of heart failure letters mentioned “memory problem,” and a mere 0.7% of letters indicated both heart failure and a memory clinic consultation. These findings contrast with cross-sectional studies reporting cognitive impairment in 25–75% of heart failure cases, raising important questions about documentation practices and the recognition of cognitive impairment in routine clinical care^2,3,7^.

The lack of listed cognitive complaints in patients with heart failure may be attributed to the lack of eliciting cognitive complaints in current heart failure diagnostic guidelines^19^. Consequently, it is likely that cognitive complaints were not inquired about or recorded during patient assessments. However, we cannot definitively determine whether patients with heart failure did not spontaneously mention cognitive complaints, explicitly denied such cognitive complaints (resulting in no records), or if healthcare providers simply did not inquire about or recognize cognitive complaints. What we can confirm is that only 2.6% of all patients with heart failure studied (146 out of the total) had "memory problem" documented in their clinical notes. In addition, our findings indicate that only 0.7% of all individuals had both heart failure and attended a memory clinic, meaning that they had both a consultation with a cardiologist and a neurologist or geriatrician. These results indicate the possibility of cognitive complaints being underrecognized in patients with heart failure in clinical practice, potentially resulting in a very limited number of patients with heart failure being referred for further cognitive assessment.

It is important to note that the previously listed prevalence of cognitive complaints in patients with heart failure (25-75%), was primarily based on questionnaires and cognitive screening rather than routinely collected healthcare data^2,3,7,20,21^. This raises the question of whether patients with heart failure might report cognitive complaints more frequently when specifically asked about them, as in targeted questionnaire studies. It is possible that when patients with heart failure visit a cardiologist in an outpatient clinic, their primary focus is on cardiac functioning and related complaints. Cognitive complaints might be perceived as unrelated to their current visit, leading patients to not mention these complaints unless specifically prompted. This absence of specific inquiry regarding cognition could explain the lower prevalence of CI-related concepts in our study compared with expectations based on cross-sectional studies^2,3,7,20,21^. However, as mentioned earlier, we cannot definitively determine whether patients with heart failure did not spontaneously mention cognitive complaints, explicitly denied these complaints (resulting in no recording), whether health care providers did not ask about cognitive complaints or did not write them down.

While the findings may indicate underrecognition of cognitive complaints, certain factors should be considered. First, although previous research cited in this study suggests a link between cognitive impairment and heart failure, cognitive impairment can arise from a range of causes, such as Alzheimer’s disease, other degenerative cerebral disorders, and chronic infectious diseases affecting the brain. Thus, the inclusion of patients attending memory clinics—who often present with mixed etiologies—may introduce variability. This reinforces the importance of interpreting the comparison between patients with heart failure and memory clinic patients as a broad contextual observation rather than a direct equivalence. Second, age-related differences between the two patient groups are an important consideration, as patients attending memory clinics are typically older than those diagnosed with HF. Since age is a significant risk factor for cognitive impairment, future studies would benefit from stratifying results by age to better account for this potential confounder.

This study has several strengths. First, this study used real-world routine care data, making the findings relevant to clinical practice. By analysing real-world clinical notes, the study provides insights into the reporting of cognitive complaints in patients with heart failure during clinical practice. MedCAT successfully extracted clinically relevant complaints for both patients with heart failure and patients attending memory clinics, in accordance with reporting guidelines for both patient groups^19,22^. Our findings underscore the feasibility of extracting medical concepts from Dutch clinical notes and analysing them using a combination of NLP-methods, specifically NER+L strategies and Association Rule Mining. Second, this study employed Named Entity Recognition and Linking (NER+L) strategies, specifically MedCAT and MedCATTrainer, to extract and categorize complaints from clinical notes. These methods identified typical complaints related to heart failure and cognition, in alignment with diagnostic guidelines.

This alignment enhances the credibility of the findings. Third, the study revealed potential underrecognition of cognitive complaints in patients with heart failure, potentially resulting in a limited number of patients with heart failure being referred for further cognitive assessment. By revealing this potential underrecognition, the study contributes to the broader awareness of the consideration of cognitive function as a routine aspect of care for patients with heart failure.

Also, several limitations should be considered. First, documentation practices, which vary from clinician to clinician, and the communication skills of the individuals may have affected the quality and availability of data and thus the complaints identified. This may lead to underreporting, which means that the prevalence of cognitive complaints in patients with heart failure may have been underestimated in our results. Second, the MedCAT algorithm had difficulty identifying complaints when they were not specifically mentioned but were implicitly embedded in a larger context. We found that the clinical notes of patients attending memory clinics were twice as long as the clinical notes of patients with heart failure. The clinical notes of patients attending memory clinics were often more comprehensive in terms of the patients’ symptom pattern; for example, the patients’ living environment was more often extensively described, in which the complaints were sometimes embedded. This may have caused a possible underestimation of the number of complaints in the clinical notes of patients attending memory clinics. The clinical notes of patients with heart failure were more often more concise, allowing the model to identify the complaints more easily in patients with heart failure, with the possible result that the number of concepts identified by MedCAT in patients with heart failure was higher and more accurate. This makes it unlikely that large numbers of cognitive complaints were missed in patients with heart failure. But that does not necessarily mean, therefore, that patients with heart failure do not experience cognitive complaints, as also described in detail before. Third, we have not yet conducted a concordance study to compare manual labelling of reports with labelling by MedCAT. This would provide additional insight into MedCAT’s model performance.

This study carries implications for clinical practice. First, it provides insights on documentation practices within routine clinical care, particularly regarding the reporting of cognitive complaints in patients with heart failure. The findings suggest that cognitive complaints are notably underlisted in clinical notes of patients with heart failure. This raises awareness of a potential gap in the recognition and management of cognitive complaints in patients with heart failure. Our study suggests the need for interdisciplinary collaboration between cardiology, neurology, and geriatrics to treat patients with heart failure. By revealing a potential underrecognition of cognitive complaints in patients with heart failure in clinical practice, it suggests that a different approach may be needed in the anamnesis of cognition in heart failure, which is supported by differences in documentation between patients attending memory clinics and patients with heart failure. Second, the study underscores the added value of employing Natural Language Processing (NLP) techniques, such as Named Entity Recognition and Linking (NER+L), in clinical research. The use of MedCAT and MedCATTrainer to extract and categorize complaints related to heart failure and cognition demonstrates the potential of NLP in analysing unstructured healthcare data. This technology can enhance the accuracy and efficiency of complaint identification in large, unstructured datasets.

This study provides opportunities for the future. First, to investigate potential information leakage at the stage between the patient’s mentioning of complaints and the clinician’s reporting, speech recognition and transcription technologies, considering privacy guidelines, could be of added value. Speech recognition and transcription could help record and automatically extract terms based on the conversations between the patient and the clinician for research purposes, making the clinician’s choices of what to note and what not to note less of a factor^23,24^. Second, more insight into screening techniques for cognitive complaints that are adequate and efficient within heart failure, so that advice on this can be given in guidelines, may be of added value.

In conclusion, this study shows a low reporting frequency of cognitive complaints in clinical notes of patients with heart failure, even though both conditions often co-occur according to cross-sectional studies. The study highlights the value of using Natural Language Processing techniques such as MedCAT and MedCATTrainer to extract complaints from unstructured electronic patient records. Opportunities for future research include investigating possible information leaks in patient-physician communication, and further exploring adequate screening techniques for cognitive complaints in heart failure that can be implemented in guidelines

## Data availability

Due to ethical and legal considerations, supporting data is not available.

## Data Availability

s we worked with electronic health records collected with an IRB waiver for informed consent from NedMec, under the disproportionate effort clause, I will not be able to share data with others.

## Supplementary Tables

**Supplementary Table S1.** Example of a concept database entry, (Dizziness) used to link complaints identified within clinical notes to biomedical databases

| Column | Description | Example values |
| --- | --- | --- |
| **CUI** | Concept id | C0012833 |
| **Name** | Term name | Duizeligheid (Dizziness) |
| **Name status** | Term type in source | PN (Primary name) |
| **Type ID** | Semantic type identifier | T047 (Sign of Symptom) |
| **Ontologies** | Source Ontology | MSHDUT, MDRDUT |

**Supplementary Table S2.** Results of Apriori algorithm to assess co-occurrence of commonly listed complaints in patients attending at memory clinics in clinical notes of patients with heart failure.

| LHS |  | RHS | Confidence | Count |
| --- | --- | --- | --- | --- |
| {Memory Problem} | => | {Get Lost} | 0.061 | 9 |
| {Dizziness} | => | {Get Lost} | 0.004 | 7 |
| {Chest Pain} | => | {Get Lost} | 0.002 | 6 |
| {Dizziness, Memory Problem} | => | {Get Lost} | 0.097 | 6 |
| {Chest Pain, Dizziness} | => | {Get Lost} | 0.005 | 6 |
| {Dizziness} | => | {Morose} | 0.008 | 14 |
| {Dyspnea} | => | {Morose} | 0.004 | 14 |
| {Memory Problem} | => | {Morose} | 0.068 | 10 |
| {Dizziness, Dyspnea} | => | {Get Lost} | 0.009 | 10 |
| {Palpitations} | => | {Morose} | 0.004 | 9 |

---

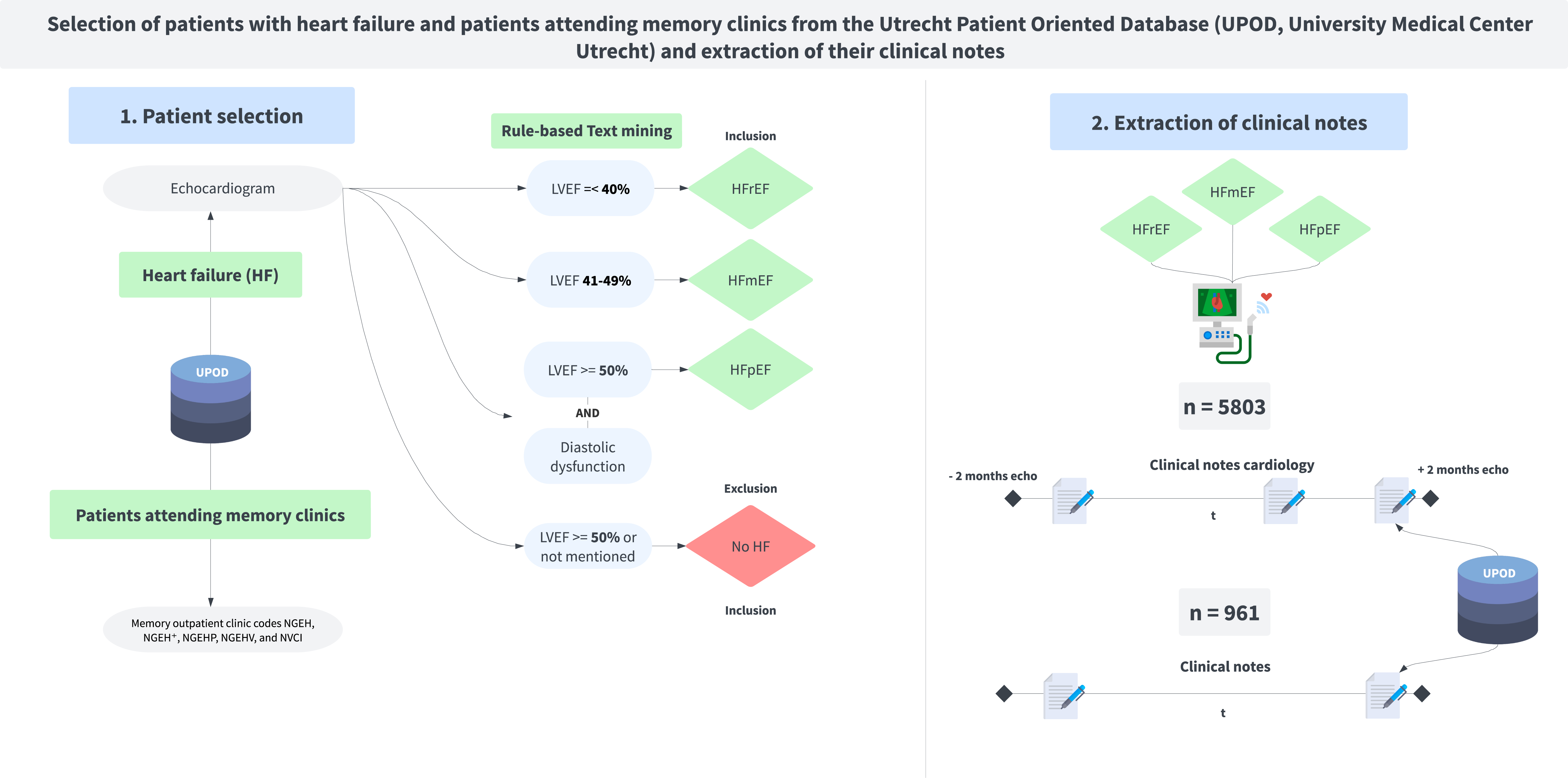

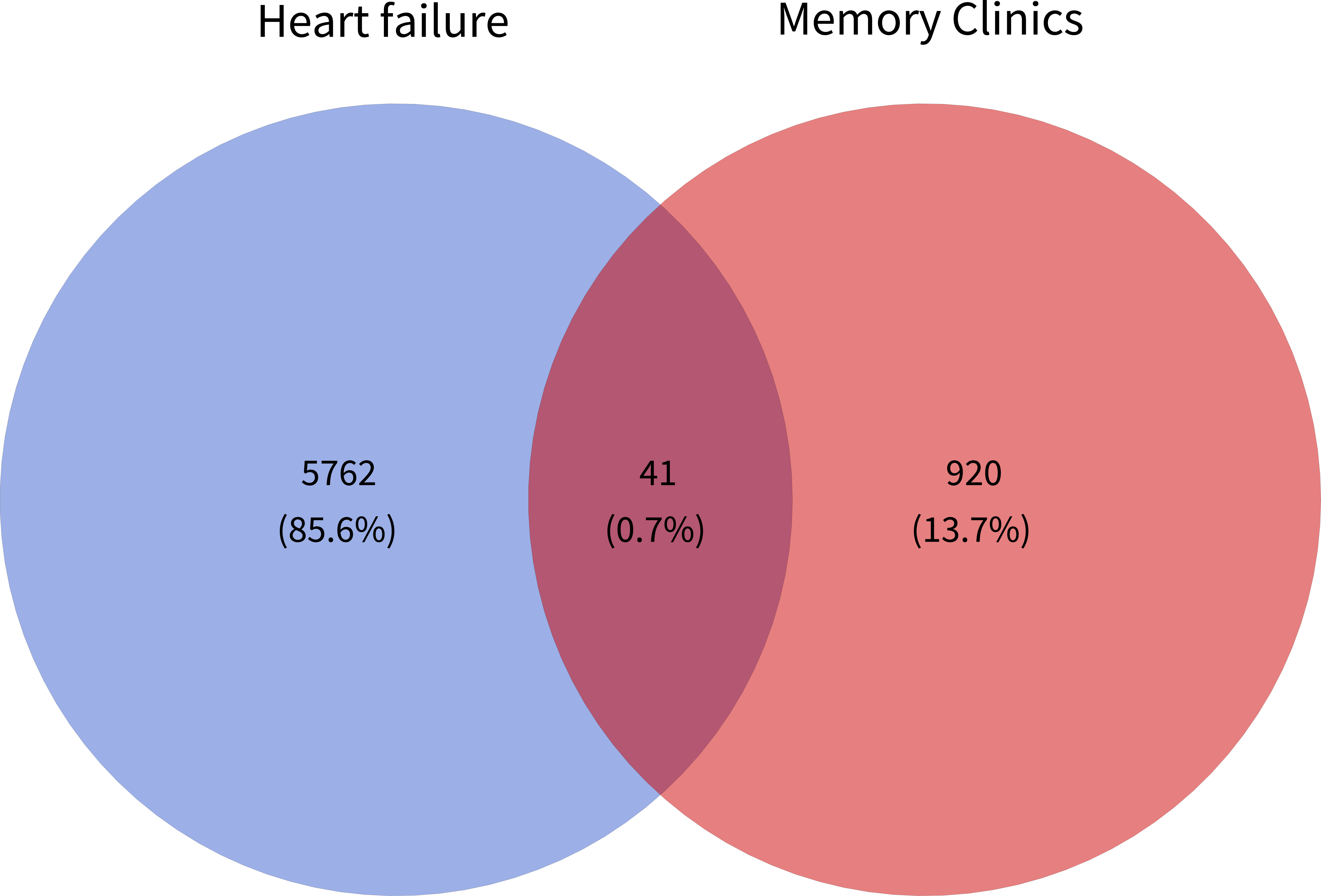

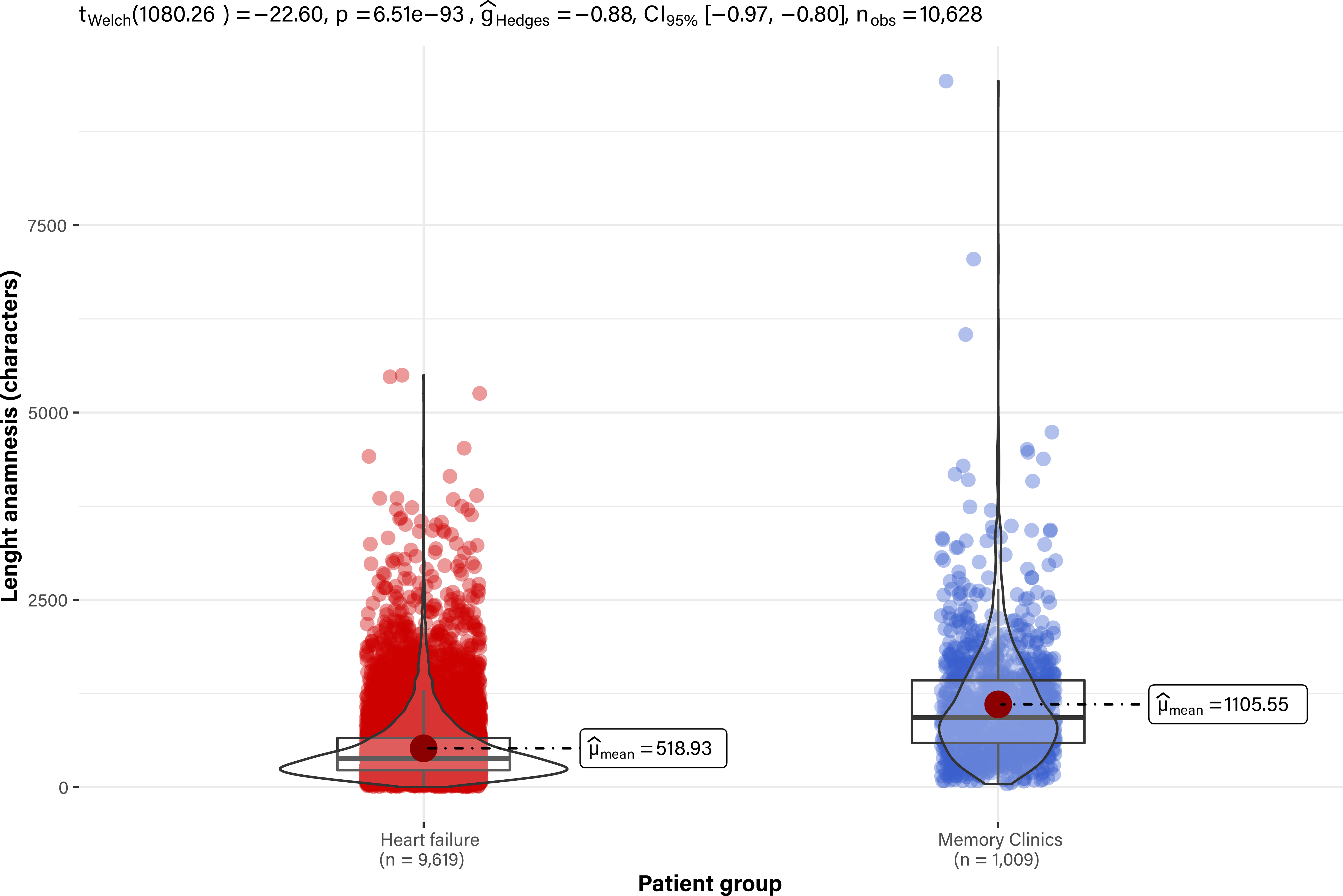

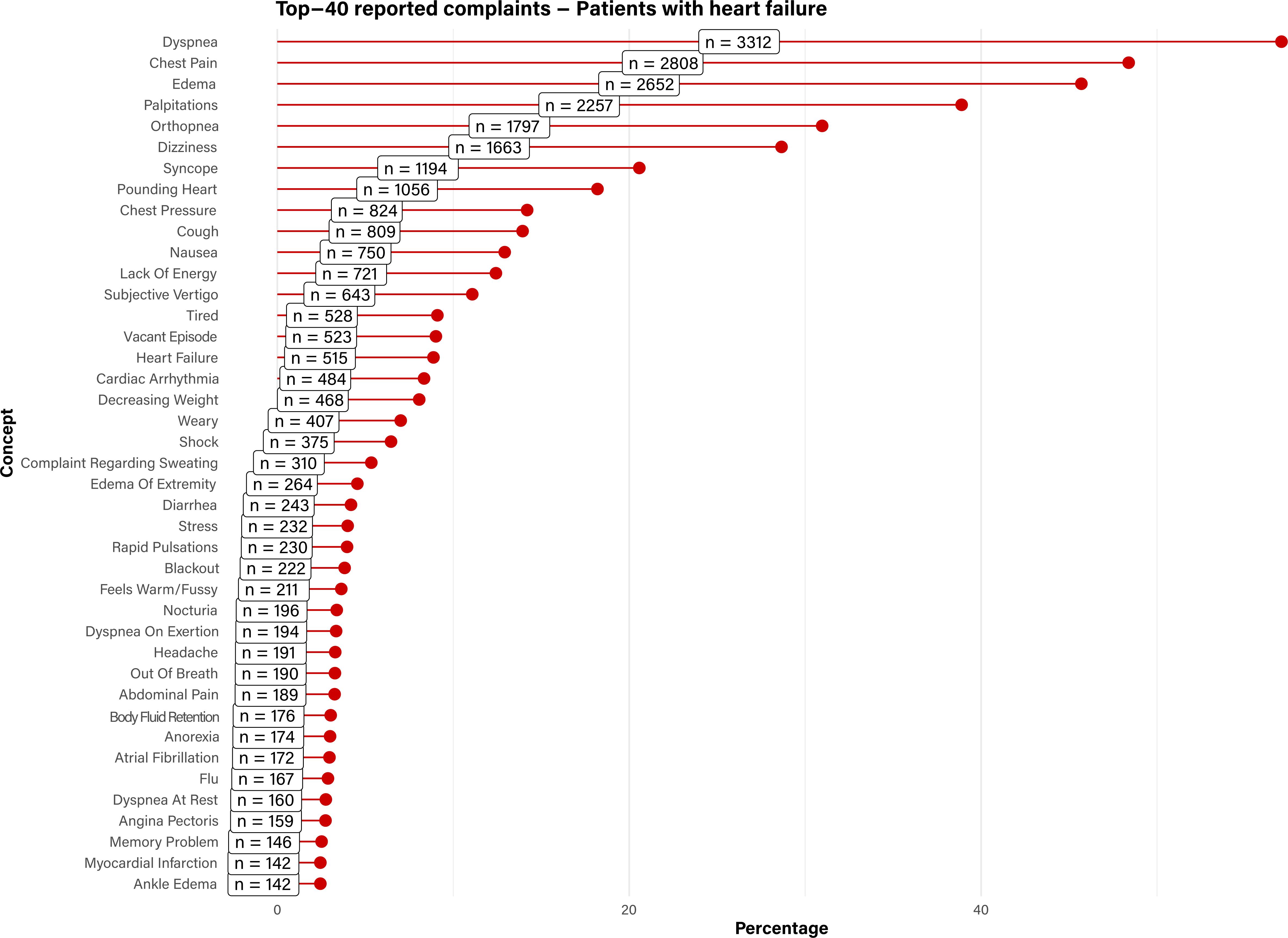

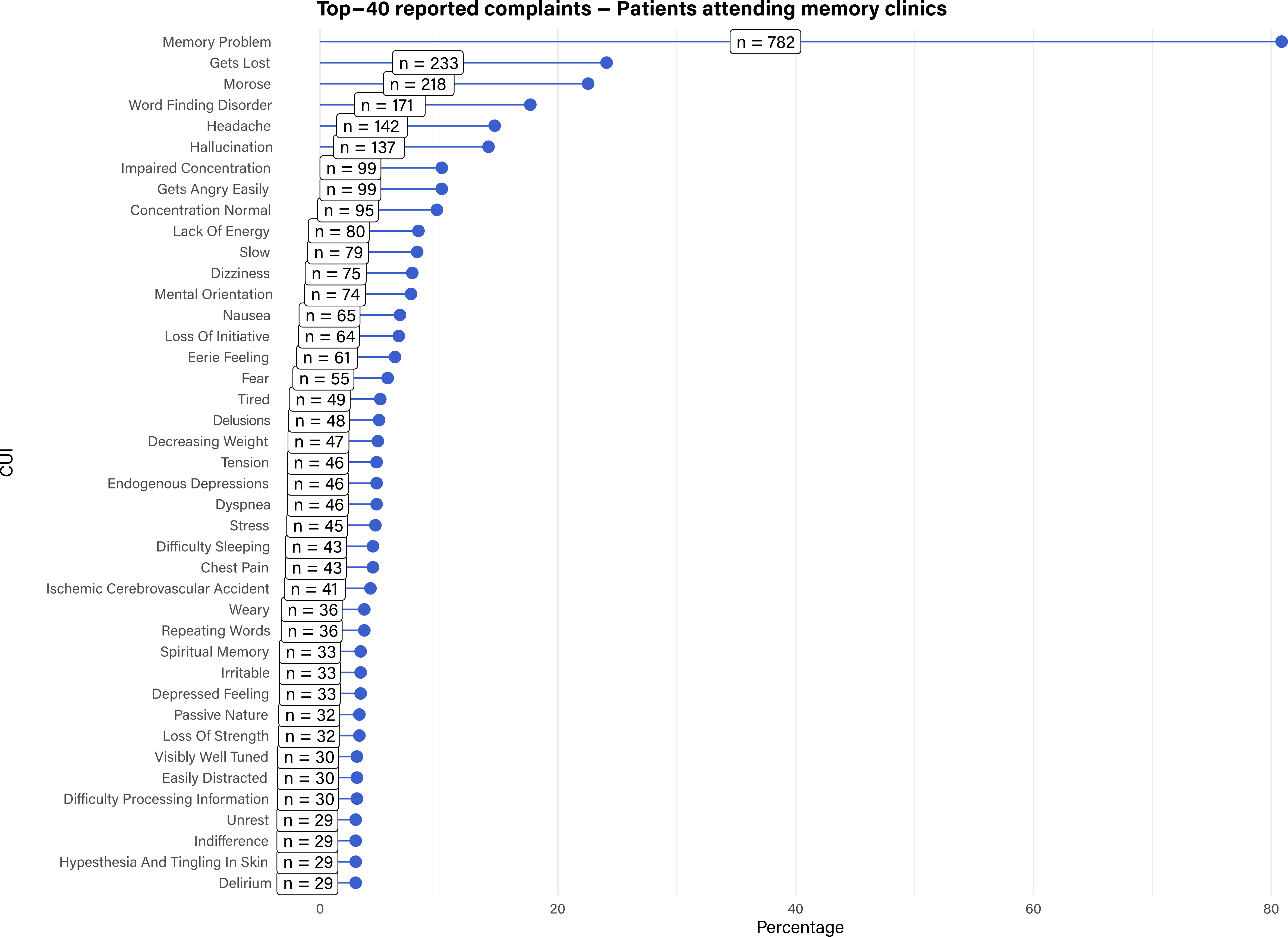

